# Behavioural barriers to COVID-19 testing in Australia

**DOI:** 10.1101/2020.09.24.20201236

**Authors:** Carissa Bonner, Carys Batcup, Julie Ayre, Kristen Pickles, Rachael Dodd, Tessa Copp, Samuel Cornell, Erin Cvejic, Thomas Dakin, Jennifer Isautier, Brooke Nickel, Kirsten McCaffery

## Abstract

**Background:** The current suppression strategy for COVID-19 in Australia is dependent on people getting tested and self-isolating while they have COVID-19 symptoms. However, there is very little research on the behaviours and behavioural barriers involved in getting tested, both in Australia and worldwide, despite there being some evidence that these barriers do exist.

**Methods:** The Sydney Health Literacy Lab (SHeLL) has been conducting a national longitudinal survey in Australia since April 2020. A list of testing barriers was included in Wave 3 in June 2020 (n=1369), along with intentions to test and self-isolate if symptomatic. Open responses were also collected. The test barriers identified were categorised using the COM-B framework.

**Results:** Only 49% of people strongly agreed they would get tested if they had COVID-19 symptoms, but most people agreed to some extent that they would get tested (96%). The most common barriers selected from the list provided were that testing is painful (11%), not knowing how to get tested (7%), and worry about getting infected at the testing centre (5%). Many participants (10%) indicated other reasons, and open responses included many additional barriers to testing than those provided in the initial list. These covered all components of the COM-B model.

**Conclusion:** We identified a wide range of barriers using both quantitative and qualitative methods, which need to be addressed in order to increase COVID-19 testing behaviour.

## BACKGROUND

The current suppression strategy for COVID-19 in Australia is dependent on people getting tested and self-isolating while they have COVID-19 symptoms (e.g. fever, cough, sore throat). According to the COM-B model, COVID-19 prevention behaviours can be conceptualised in terms of three main drivers: physical/psychological *capability* (e.g. having the physical ability to drive to and walk up the stairs to access a testing centre, and knowing what to do if you get a cough), physical/social *opportunity* (e.g. the availability of testing centres in your area, and social norms that make self-isolation acceptable), and automatic/reflective *motivation* (e.g. getting into the habit of staying home when you have symptoms, and believing that it ‘s important to get tested if you develop symptoms)^1^. Furthermore, a person who is already motivated to test will require different strategies (e.g. to overcome specific access barriers) than a person who does not see the value in testing and is not motivated to test. For example, the Health Action Process Approach suggests that intentions are driven by risk perception about the threat, self efficacy to take action to reduce the threat, and what you expect the outcomes of the threat and action will be^2^. But to translate intentions into behaviours, you also need to plan exactly what you will do (action planning) and how you will address barriers to that plan (coping planning)^2^.

There is little research on COVID-19 testing behaviours given the very new nature of this issue, but current literature and media reports suggest different barriers exist across countries. For example, countries such as Tanzania have major issues with opportunity in terms of limited access and fake testing kits^3^. Cost may be a barrier in other countries such as the US where health insurance may not cover the testing. This disproportionately affects certain groups such as immigrant and non-citizen communities, who may also fear financial and legal repercussions from testing positive^4^. Furthermore, testing may be limited to certain criteria (e.g. only if you have symptoms, regardless of exposure to COVID-19 cases) due to lack of supply or staff resource issues^5^. There may also be issues of delivering tests and transporting samples for remote areas^6^. Finally, there may be issues with inadequate communication and low community knowledge about which symptoms require testing and the process to follow^5^.

Australia is fortunate to have efficient and freely accessible testing widely available, although this can vary by location. Testing clinics have been set up all around the country including drive through options to minimise contact with other patients, and results are generally sent by text message within 2 days^7,8^. However, the process can take longer and is unpredictable where demand increases in new outbreak areas such as Victoria in July-September 2020^9–11^.

We know that certain groups are less likely to understand and act on COVID-19 prevention advice. For example, people with lower health literacy and those who speak a language other than English were less likely to be able to identify COVID-19 symptoms and prevention measures^12^. Younger people, men and those with less education were more likely to agree with COVID-10 misinformation about prevention, management and cure^13^. These groups likely need different communication strategies to ensure everyone *understands* the message and takes appropriate *action* based on the message.

Messaging trials have tested different strategies to improve intentions to follow COVID-19 prevention advice, for example framing the motivation as helping the community rather than avoiding individual risk^14^, or testing identity-based messages such as “don ‘t be a spreader”^15^. However, emerging research in this area is generally not peer reviewed and has neglected testing as a key behavioural outcome for COVID-19 prevention. We need to understand the behavioural barriers^1^ to testing in order to develop and evaluate interventions to address these issues.

## AIM

This study aims to address a major gap in our understanding about how to improve COVID-19 testing behaviour, by: 1) reporting the prevalence of specific test barriers in an Australian survey sample; 2) identifying additional test barriers through open responses, since research on this is still emerging; and 3) linking these barriers to an overarching framework of behaviour change.

## METHOD

### National survey

The Sydney Health Literacy Lab (SHeLL) has been conducting a national longitudinal survey in Australia since April 2020^12,13^. The original sample was recruited via an online market research panel, supplemented with social media advertising (n=4326). The social media users were followed-up monthly from April-July. A list of testing barriers of interest to the NSW Department of Health was included in Wave 3 in June 2020 (n=1369), along with intentions to test and self-isolate if symptomatic. These were worded as: “over the next 4 weeks, I plan to get tested if I have COVID-19 symptoms (cough, sore throat, fever)” and “over the next 4 weeks, I plan to stay home if I have COVID-19 symptoms (cough, sore throat, fever)”; with 1-7 response options from strongly disagree to strongly agree. Open responses to “other” test barriers were also collected. Descriptive analyses are reported as percentages for the survey results.

### COM-B behavioural diagnosis

The test barriers identified from the survey responses were categorised using the COM-B framework^16^ by 2 researchers who have been trained in the use of this framework (CB and CAB). CAB conducted the initial coding, and this was reviewed by CB with discussion of issues that could be coded in different ways until consensus was reached.

## RESULTS

The results of the survey questions are reported in Tables 1-2. Only 49% of people strongly agreed they would get tested if they had COVID-19 symptoms (cough, fever, sore throat), but most people agreed to some extent that they would get tested (96%). For self-isolation, 69% strongly agreed they would stay home if they had symptoms. The most common barriers selected from the list provided were that testing is painful (11%), not knowing how to get tested (7%), and worry about getting infected at the testing centre (5%). Many participants (10%) indicated other reasons, and open responses included many additional barriers to testing than those provided in the initial list. Table 3 maps all the barriers identified in open survey responses to the COM-B drivers of behaviour, covering all components.

**Table 1:** Intentions if symptomatic for COVID-19 over the next 4 weeks.

| **Intention to get tested if symptomatic in next 4 weeks** | **n** | **%** |
| --- | --- | --- |
| Strongly agree | 674 | 49.1 |
| Agree | 376 | 27.4 |
| Somewhat agree | 101 | 7.4 |
| Neither agree nor disagree | 72 | 5.2 |
| Somewhat disagree | 40 | 2.9 |
| Disagree | 49 | 3.6 |
| Strongly disagree | 60 | 4.4 |
| **Intention to self-isolate if symptomatic in next 4 weeks** | **n** | **%** |
| Strongly agree | 951 | 69.3 |
| Agree | 310 | 22.6 |
| Somewhat agree | 53 | 3.9 |
| Neither agree nor disagree | 12 | 0.9 |
| Somewhat disagree | 14 | 1.0 |
| Disagree | 6 | 0.4 |
| Strongly disagree | 26 | 1.9 |

**Table 2:** Barriers to testing if symptomatic for COVID-19.

| **Testing barrier** | **n** | **%** |
| --- | --- | --- |
| Testing is painful | 153 | 11.2 |
| Other (please detail) – see Table 3 | 136 | 9.9 |
| I don't know how, when and where to get tested | 98 | 7.1 |
| I'm worried I will get infected with COVID-19 at the testing clinic | 81 | 5.9 |
| I'll forget to get tested | 33 | 2.4 |
| I'm worried about what others think of me | 32 | 2.3 |
| It's too difficult or expensive to get tested | 29 | 2.1 |
| Testing doesn't work / I don't trust the results | 17 | 1.2 |
| No one else is getting tested | 11 | 0.8 |

**Table 3:**
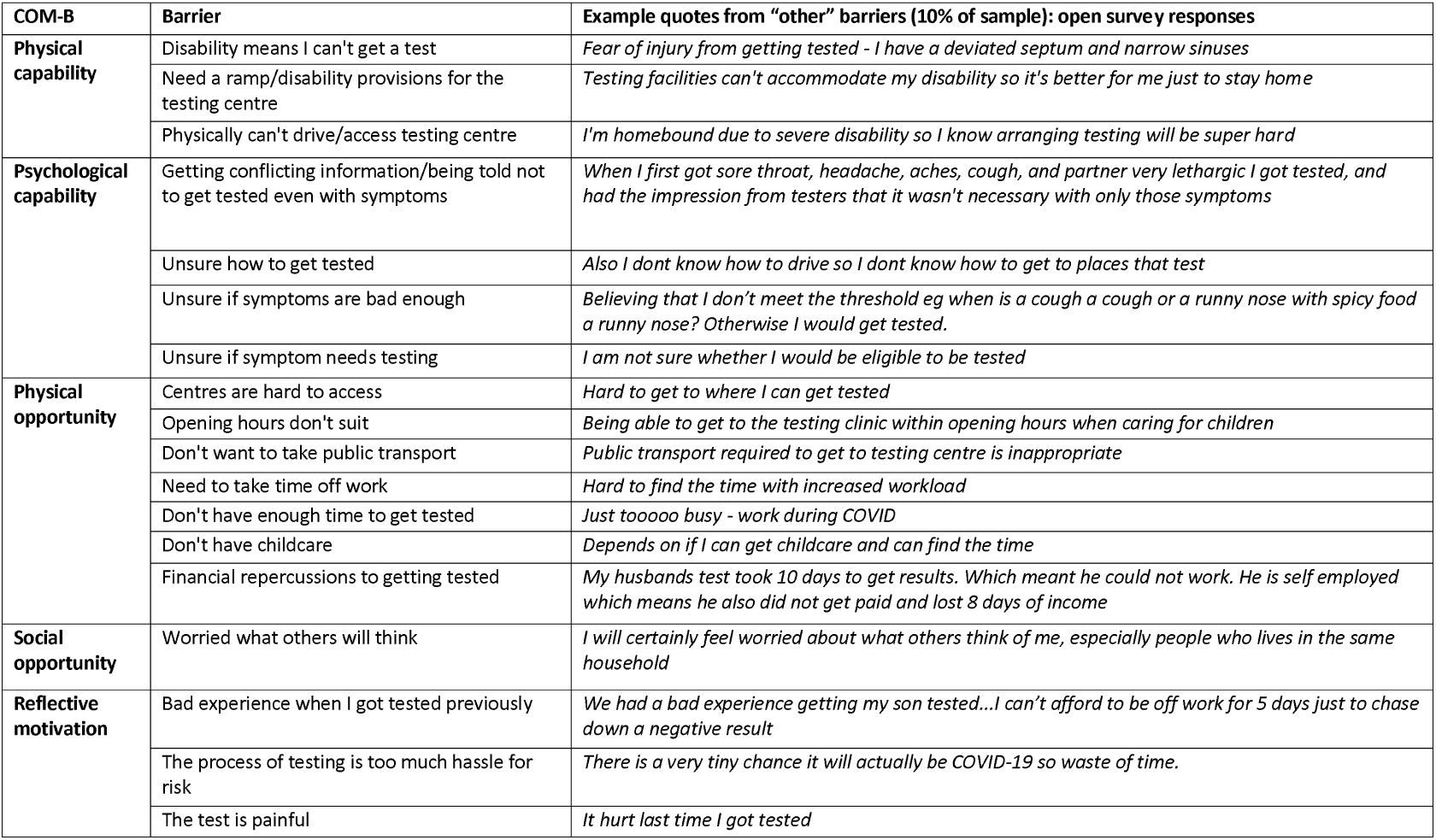
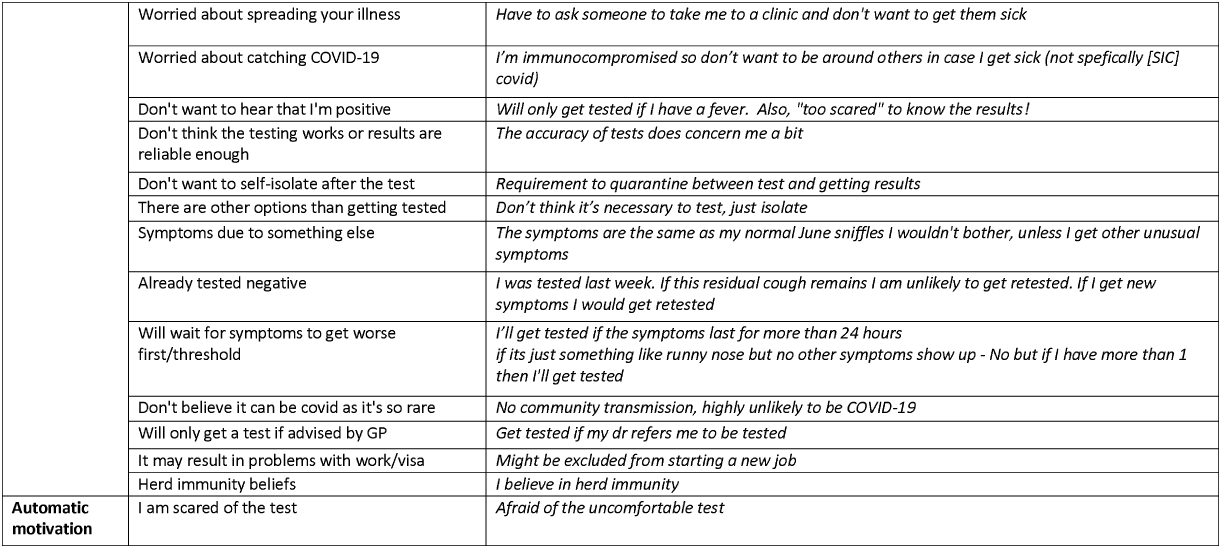
COM-B drivers of COVID-19 testing behaviour.

## DISCUSSION

This paper provides insights into testing barriers in Australia, showing that they cover all possible theoretical drivers of behaviour: capability, opportunity and motivation. Since this is such a complex issue, it requires a multifaceted approach to address.

For capability barriers, we know that government information about COVID-19 is difficult to understand for the average person based on readability analyses^17^, and that communities with different language needs are not being adequately addressed^18^. We have also identified that certain groups are more sucsceptible to misinformation, such as younger people and men^13^. Addressing different information needs in the community may require using less traditional news sources^19^ and different spokespeople to reach these groups, such as social media influencers^20^.

Although the Australian testing system is comparatively accessible, there are still opportunity barriers particularly around financial losses and work expectations that require system changes. Victoria has addressed this during the recent outbreak by providing financial support for those who are self isolating^21^. There are also social opportunity issues where stigma and perceived judgment by others could prevent individuals from getting tested, which we have seen in other infectious disease conditions such as HIV^22^.

The largest category of barriers in this study was motivation, which is likely to influence both intentions and behaviours. To address low intentions to test we need to identify and address the specific concerns that each individual has, as they are so wide ranging. For some people, online symptom checkers could be a channel to provide tailored information, where users could be prompted for key concerns to link them to the most relevant information^23,24^, as opposed to searching for this in long FAQ pages on government health websites. However, they are not all reliable^25^, and may not be accessed by some groups such as culturally and linguistically diverse communities. To address gaps between intentions and actual behaviour, we could help people to plan in advance for the inconvenience of testing and self isolation, for example a person could have a plan worked out in advance with their manager for how they will notify work and change shifts or work from home. Setting goals in advance has been shown to change various health-related behaviours^26,27^, and this strategy has been applied in Australia to help communities plan for bushfire management^28^.

Our findings from June are very similar to September results from national flu tracking data, which show 51% of people who experienced COVID-19 symptoms did not get tested^29^. Our findings show 51% of people did not “strongly agree” they would get tested, but most people agreed to some extent and addressing barriers may improve this overall positive orientation and address the behaviour gap. Another survey conducted in August found much higher rates of testing avoidance, and reported 85% of respondents who had sympoms had not been tested^30^.

In conclusion, we identified a wide range of barriers using both quantitative and qualitative methods, which need to be addressed in order to increase testing behaviour. Interventions to improve testing uptake need to identify key barriers for target populations, and specifically address these using evidence-based behaviour change techniques. Our findings support broader advice from international experts in behaviour change, highlighting the importance of diagnosing behavioural barriers in order to increase compliance with COVID-19 prevention behaviours^1^.

## Data Availability

The data is stored in a password-protected server at the University of Sydney

## ACKNOWLEDGEMENTS

We thank the participants of the longitudinal COVID-19 survey for their ongoing participation in this research. This study was not specifically funded, but in-kind support was provided by authors with research fellowships. CB is supported by a National Health and Medical Research Council (NHMRC)/Heart Foundation Early Career Fellowship (#1122788).

RD is supported by a University of Sydney fellowship (#197589).

KM is supported by a National Health and Medical Research Council (NHMRC) Principal Research Fellowship (#1121110).

